# Development of a Claims-Based Computable Phenotype for Ulcerative Colitis Flares

**DOI:** 10.1101/2025.01.26.25321138

**Authors:** Daniel Copeland, Jayson S. Marwaha, Daniel Wong, William Yuan, Michelle N. Fakler, Chris J. Kennedy, Brendin Beaulieu-Jones, Vitaliy Poylin, Joseph Feuerstein, Gabriel A. Brat

## Abstract

**Background and Aims:** Several conditions exist that do not have their own unique diagnosis code in widely-used clinical terminologies, making them difficult to track and study. Acute severe ulcerative colitis (ASUC) is one such condition. There is no automated method to identify patients admitted for ASUC from observational data, nor any specific billing or diagnosis code for ASUC. Accurate, automated, large-scale identification of hospital admissions for non-coded conditions like ASUC may enable further research into them.

**Methods:** We performed a retrospective cohort study of patients with a history of ulcerative colitis (UC) admitted to a single academic institution from 2014-2019. Clinicians at our institution performed a chart review of these admissions to determine if each was due to a true episode of ASUC or not. Logistic regression, random forest (RF), and support vector machine (SVM) models were trained upon administrative claims data for all admissions.

**Results:** 268 ASUC admissions and 3,725 non-ASUC admissions among UC patients were included. Our RF model exhibited the best performance, correctly classifying 95.5% of admissions as either ASUC or non-ASUC, with a validation AUROC of 0.96 (95% CI 0.94-0.98; AUPRC 0.73). The model had a sensitivity of 81.5% and specificity of 96.5%. The five most important features in the model were endoscopy of sigmoid colon, length of stay, age, endoscopy of rectum, and abdominal x-ray.

**Conclusions:** There is currently no modality by which ASUC, which does not have its own unique diagnosis code, can be identified from claims databases in a scalable fashion for research or clinical purposes. We have developed a machine learning-based model that identifies clinically significant ASUC and reliably distinguishes them from admissions for non-ASUC reasons among UC patients. The ability to automatically curate large, accurate datasets of non-coded conditions like ASUC episodes can serve as the basis of large-scale analyses to maximize our ability to learn from real-world data, enable future research, and better understand these diseases.

**Summary:** There is currently no accurate way to identify, track, or study acute severe ulcerative colitis (ASUC) using administrative claims datasets. We have built a machine learning model to identify ASUC from claims data to enable large-scale studies on this condition.

## Introduction

Several medical conditions exist that do not have their own unique diagnosis code in widely-used clinical terminologies.^1^ This is a significant limitation of real-world clinical data: conditions that do not have a unique identifier can be difficult to pick out from structured observational data sources such as administrative claims databases. These non-coded conditions, as a result, cannot be easily tracked for clinical purposes or studied for research purposes, despite having physician-defined characteristics.^2^ The lack of identifiers for non-coded conditions that are poorly understood is a significant barrier to improving our understanding of them, since large datasets of these conditions cannot be easily curated to enable study into their risk factors, clinical trajectories, or pathophysiology.^3^ Several examples of such non-coded conditions exist;^4^ in this study, we focus on acute severe ulcerative colitis (ASUC).

Ulcerative colitis (UC) is a chronic, debilitating, inflammatory bowel disease characterized by periods of remission with few symptoms interspersed with periods of acute exacerbation. The American Gastroenterology Association (AGA) defines varying degrees of disease activity with ASUC being the most severe manifestation of UC. ASUC is an exacerbation of moderate-to-severe UC that requires hospitalization and is characterized by having six or more bloody bowel movements per day and at least one marker of systemic toxicity.^5^ This is a highly morbid condition: approximately one third of patients experiencing ASUC will require a colectomy within one year.^6,7^ Due to this morbidity and the high cost of patients enduring ASUC, a better understanding of drivers of this condition would be of great value. The ability to identify ASUC episodes from national retrospective data to power large observational studies is an essential - but missing - first step in improving our understanding.

The risk factors of UC exacerbations, including ASUC, are multifactorial and poorly understood. Potential triggers include adverse response/failure to respond to treatments, abnormal immune response, genetics, microbiome, stress and environmental factors.^5,8^ Additionally, the grading of UC disease activity often relies on subjective information including pain, characterization of bowel movements, and endoscopic impression of the colon.^7,9–11^ Due to their multifactorial and subjective nature, ASUC episodes have proven very difficult to prevent or treat.

There is currently no method to identify patients admitted for ASUC from large administrative claims databases, nor is there a specific billing code to denote an admission related to ASUC. The ability to identify ASUC admissions easily from administrative billing data could potentially serve as a very valuable tool to unlocking large-scale analyses and better understanding this condition. Prior studies that attempt to study UC exacerbations using administrative data have independently generated several different code-based rules for defining flares.^12–14^ The true performance of these different code-based definitions is unknown, calling into question the accuracy of the cohorts they studied. Importantly, these rule-based methods have been developed without first understanding if there is sufficient data in administrative claims databases to make the diagnosis confidently, what best methods should be used, or what best predictors exist in this data that should be leveraged. The incompleteness of administrative claims databases is well-documented phenomenon^15^ and necessitates the need for a more flexible machine learning solution that learns patterns robust to code missingness, rather than rule-based methods that depend on specific codes that may have a high likelihood of missingness. There are limited large-scale studies of UC flares for this reason.

The purpose of this study is to develop a machine learning-based methodology for automatically identifying patients with ASUC that are not explicitly coded within large-scale structured observational data sources such as administrative and insurance claims databases that typically lack details about patients that are granular enough for a human to make a confident diagnosis. Instead of using an administrative code, we use physician-defined characteristics (i.e., the AGA-defined clinical characteristics of ASUC) to diagnose the condition in the EHR, then train a machine learning classifier to identify patterns in administrative claims data that corresponds to this EHR-based diagnosis. The ultimate goal of our work is to be able to generate large datasets of ASUC from nationwide claims datasets with limited patient-level data, which then can be used in the future to better understand the pathophysiology and risk factors for this condition, enable future research on it, and train clinically-impactful predictive tools for it.

## Materials and Methods

### Study Design

We conducted a retrospective cohort study of all UC patients who were admitted to our single academic medical center for any reason. Admissions from January 1, 2014 to June 31, 2019 were included. The study was approved by the Institutional Review Board (IRB) of Beth Israel Deaconess Medical Center (BIDMC). Physicians used the EHR data from each patient (notes, lab values, etc.) to manually label each admission as flare/no flare. We then used these labels, combined with only claims data from these patients, to train and internally validate machine learning models to classify admissions as flare or not.

### Patient Cohort

All patients 18 years and older admitted to inpatient status due to an episode of ASUC were included. The diagnostic criteria for ASUC episodes is described in further detail below. Patients were excluded if they had a history of identified colonic malignancy or colon resection at the time of admission in order to eliminate from our dataset any cases of pouchitis or postoperative complications that may have been easily mistaken for ASUC. A CONSORT-style diagram showing the full data flow of our study can be found in **Figure 1**.

**Figure 1.**
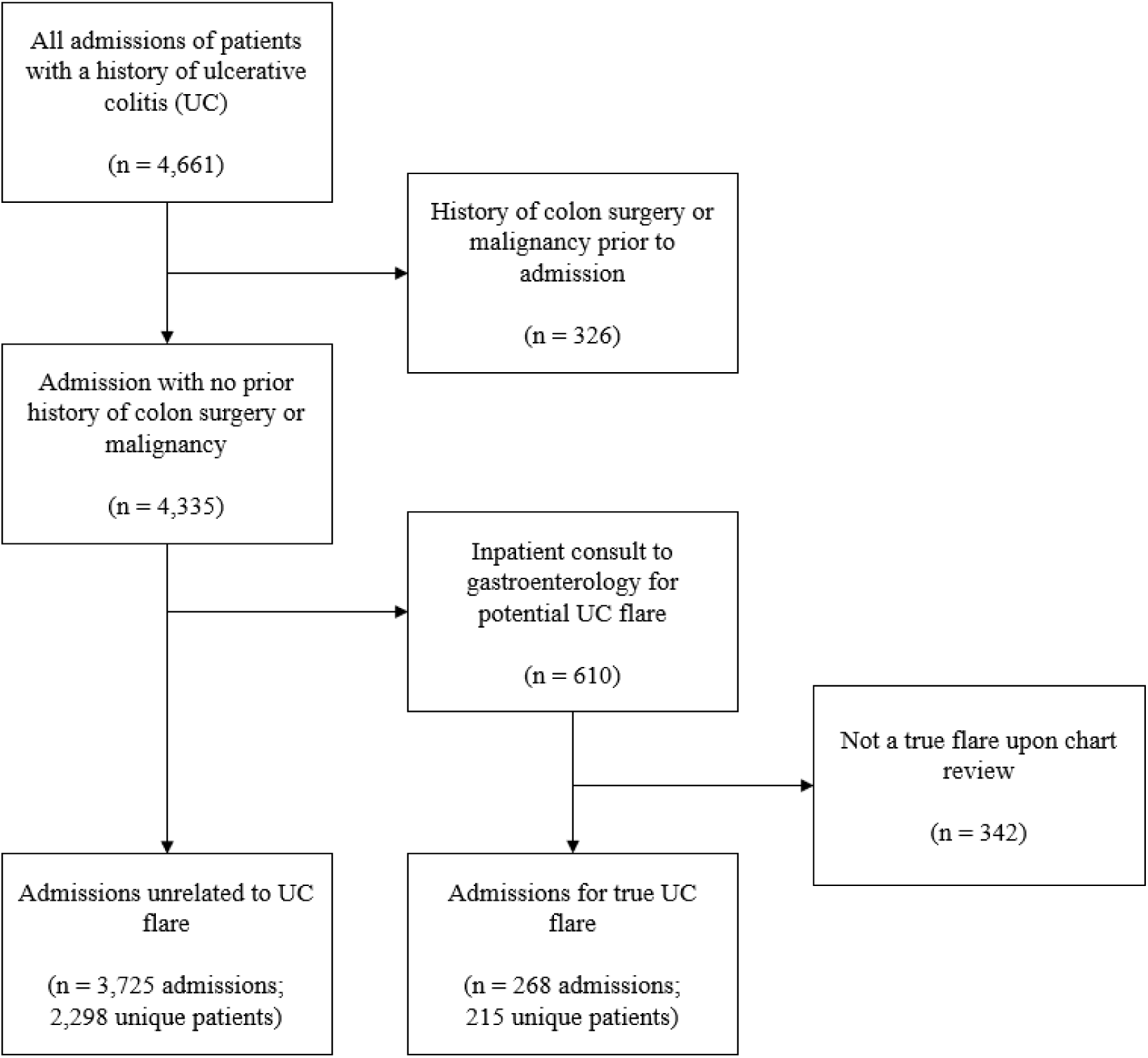
Data flow diagram.

### Data Collection

#### Identification of Billing Codes

An institutional administrative claims registry was queried for the list of admitted UC patients, defined as inpatient admissions of patients with at least 1 ICD-9 or ICD-10 diagnosis code for UC in their history during the study period. For each admission, all claims codes were extracted from the registry including ICD-9 and -10 diagnosis and procedure codes, Current Procedural Terminology® (CPT) procedure codes, and RxNorm drug codes. Dates of admission and discharge and patient demographic information were also collected. To standardize diagnosis codes across the dataset, a Unified Medical Language System (UMLS)-based crosswalk table was used to convert ICD-9 codes to their ICD-10 equivalent. In cases where ICD-9 codes mapped to multiple ICD-10 codes, we manually chose the least granular ICD-10 code.

#### Curation of ASUC Dataset

Inpatient consultations to the gastroenterology service at our institution were used to help identify admissions due to ASUC. One of the authors (JF) maintains a registry of all gastroenterology service consults for UC. We included the records of all UC patients who were admitted and seen as a consult for possible ASUC from January 1, 2014 to June 30, 2019. This time period was chosen to best resemble contemporary care including routine laboratory and endoscopic evaluation and initiation of biologic therapy in the acute setting.

Labels generated by the on-service gastroenterology fellow who originally saw the consult were confirmed by manual physician review of all charts, performed by three of the authors (DW, MF, JDF). Each admission was labeled as an ASUC or non-ASUC admission (i.e., a UC patient who was admitted for reasons unrelated to ASUC). Admissions were labelled as ASUC if there was a note from the gastroenterologist specifying that the patient had a severe exacerbation of symptoms attributable to UC, there was no documentation of a confounding diagnosis (i.e., no prior colectomy or readmission for a chronic flare), no documented uncertainty about the diagnosis (i.e. no diagnosis of colitis of unknown etiology or colitis not otherwise specified), and the patient’s clinical data was consistent with ASUC. Patients who were admitted with a high suspicion for ASUC and empirically treated with IV steroids on the day of admission - but discontinued when an alternate diagnosis was identified - were also excluded during physician review. The resulting dataset was a list of true ASUC episodes with inpatient admission. ASUC episodes were not graded with further granularity using Truelove and Witts Criteria^16^ or any other severity index due to insufficient detail in some patient charts; some notes were missing required elements of these severity grading tools.

Non-ASUC admissions among patients with a history of UC served as our control group. This control group would train the machine learning algorithm to classify between two very similar groups, patients admitted for ASUC versus patients with UC who were admitted for any other reason.

### Model Development and Validation

#### Feature Selection

To decrease the number of codes for model training, we reduced the dimensionality of the dataset using the Boruta algorithm.^17^ The Boruta algorithm selects significant features by creating “shadow” features from shuffled real features. Each real feature and its shadow features are then used to construct a random forest model, and the importance of each feature is measured. A real feature is considered significant (i.e., useful to the model) and thus included in the final model only if it outperforms all shadow features of itself.

#### Development and Validation

Logistic regression, random forest (RF), and support vector machine (SVM) models were used to predict the outcome of interest and uncover significant codes related to identifying ASUC. The outcome of interest was admission for ASUC vs. admission for non-ASUC diagnosis. To mitigate class imbalance due to the relative infrequency of ASUC admissions, we up-sampled our dataset using sampling with replacement to make class distributions equal prior to training our models. Data were randomly divided into training and test sets with an 80:20 split. Logistic regression, RF, and SVM models were trained and validated using 10-fold stratified cross-validation.

#### Relative Feature Importance

Following construction of our RF model, we used out-of-bag (OOB) error measurements to estimate the relative ranking of feature importance. The model’s OOB error was calculated with the individual removal of each feature. The larger the increase in RF OOB error when a feature is removed, the greater that feature’s importance is to the model’s prediction accuracy. We developed, validated, and reported the model in full compliance with the Equator Network Transparent Reporting of a Multivariable Prediction Model for Individual Prognosis or Diagnosis (TRIPOD) guidelines (**Supplementary Table S1**).^18^ All data analysis was performed using R version 3.6.3.

## Results

A query for all patients with a history of UC from our institutional database returned 6,108 patients. These patients were admitted to the hospital for any reason 4,661 times from 2014 to 2019. After excluding all admissions in which the patient had a history of colon surgery or colonic malignancy (n=326), 4,335 admissions were included. During 610 of these admissions, an inpatient consult was placed to the gastroenterology service for a potential ASUC episode. Upon manual physician review of all of these admissions, 342 of these admissions were determined to not be a true ASUC episode and thus were excluded. 268 admissions were determined to be due to true ASUC episodes, representing 215 unique patients. The remaining 3,725 admissions, representing 2,298 unique patients, were for a reason unrelated to ASUC and served as our control group. Data flow and exclusion criteria are described in **Figure 1**.

Demographics of the patients included in our study and the characteristics of ASUC and non-ASUC admissions are shown in **Table 1**. Patients admitted for a non-ASUC related reason were slightly older (53.8y vs. 45.2y) but the distribution of sex between the two groups was similar (56.0% female vs. 55.3% female). Patients were largely similar between the two groups, one notable exception being that patient admitted for a flare had the UC diagnosis code K51 present at significantly higher rates than patients with a diagnosis of UC but admitted for non-flare reasons (57 vs. 23%, p<0.001).

**Table 1:** Demographic and clinical characteristics of patients with flare verus non-flare diagnosis.

| Characteristic | Admitted for Flare | Admitted for Non-Flare Diagnosis | *P* |
| --- | --- | --- | --- |
| *N* | 268 | 2,298 | - |
| Age, Mean (SD) | 45.2 (16.0) | 53.8 (16.3) | <0.001 |
| Sex |  |  | - |
| Female, No. (%) | 150 (56.0) | 1,271 (55.3) | 0.1 |
| Male, No. (%) [Ref.] | 118 (44.0) | 1,027 (44.7) | - |
| LOS, days (SD) | 6.8 (6.056) | 5.5 (7.87) | 0.05 |
| Total No. Codes (SD) | 27.4 (20.2) | 31.7 (19.7) | 0.02 |
| Received lower GI endoscopy, No. (%) | 67 (25.0%) | 275 (12.0%) | <0.001 |
| Received systemic steroids, No. (%) | 9 (3.35%) | 53 (2.3%) | 0.29 |
| Received infliximab, No. (%) | 5 (1.87%) | 12 (0.522%) | 0.01 |
| Received vedolizumab, No. (%) | 0 (0%) | 0 (0%) | - |
| Received cyclosporine, No. (%) | 1 (.37%) | 3 (.13%) | 0.34 |
| Received colectomy, No. (%) | 1 (.37%) | 0 (0%) | 0.003 |
| UC K51 Code, No. (%) | 154 (57.5%) | 527 (23.0%) | <0.001 |

The resulting dataset contained 127,064 total administrative codes and 2,427 unique codes. The Boruta algorithm was used to identify which of these 2,427 features were significant. 29 features were found to be significant. While these 29 codes only represented 1% of unique codes, they accounted for 16,993, or 13%, of the total codes in our dataset. The relative importance of these codes is shown in **Supplementary Figure S1**.

To determine the optimal number of trees in our RF model, a series of RFs containing 0-500 trees were constructed and the OOB error was calculated for each. For our model, the optimal number of trees and randomly selected variables used to build each tree was determined to be 300 and 4, respectively. Parameter tuning for our RF model is shown in **Supplementary Figure S2**.

A RF model with these parameters was trained and validated on our dataset. The model correctly classified 95.5% of the test admissions as either ASUC or non-ASUC, with a validation AUROC of 0.96 (95% CI 0.94-0.98) (**Figure 2a**) and a validation AUPRC of 0.73 (**Figure 2b**). A confusion matrix of our model applied to an internal validation dataset is shown in **Table 2**. The RF model had a sensitivity of 81.5%, specificity of 96.5%, and OOB error of 5.3%. A cutoff threshold of 0.2 was used to optimize for sensitivity. If an admission was flagged by our model, the odds ratio (OR) of being a true ASUC admission was approximately 46. A manual chart review of the 26 false positives flagged by our model in this internal validation dataset revealed that 13 (50%) of them were indeed admitted for true severe UC exacerbations as well, but these exacerbations did not precisely fit the predefined criteria of ASUC (**Table 2**). The remaining 13 patients were admitted for reasons unrelated to UC exacerbation. A multidimensional scaling (MDS) plot is shown in **Figure 3**, demonstrating that the RF model trained upon administrative claims data alone finds similarity amongst most ASUC admissions and is able to reliably separate ASUC from non-ASUC admissions.

**Figure 2.**
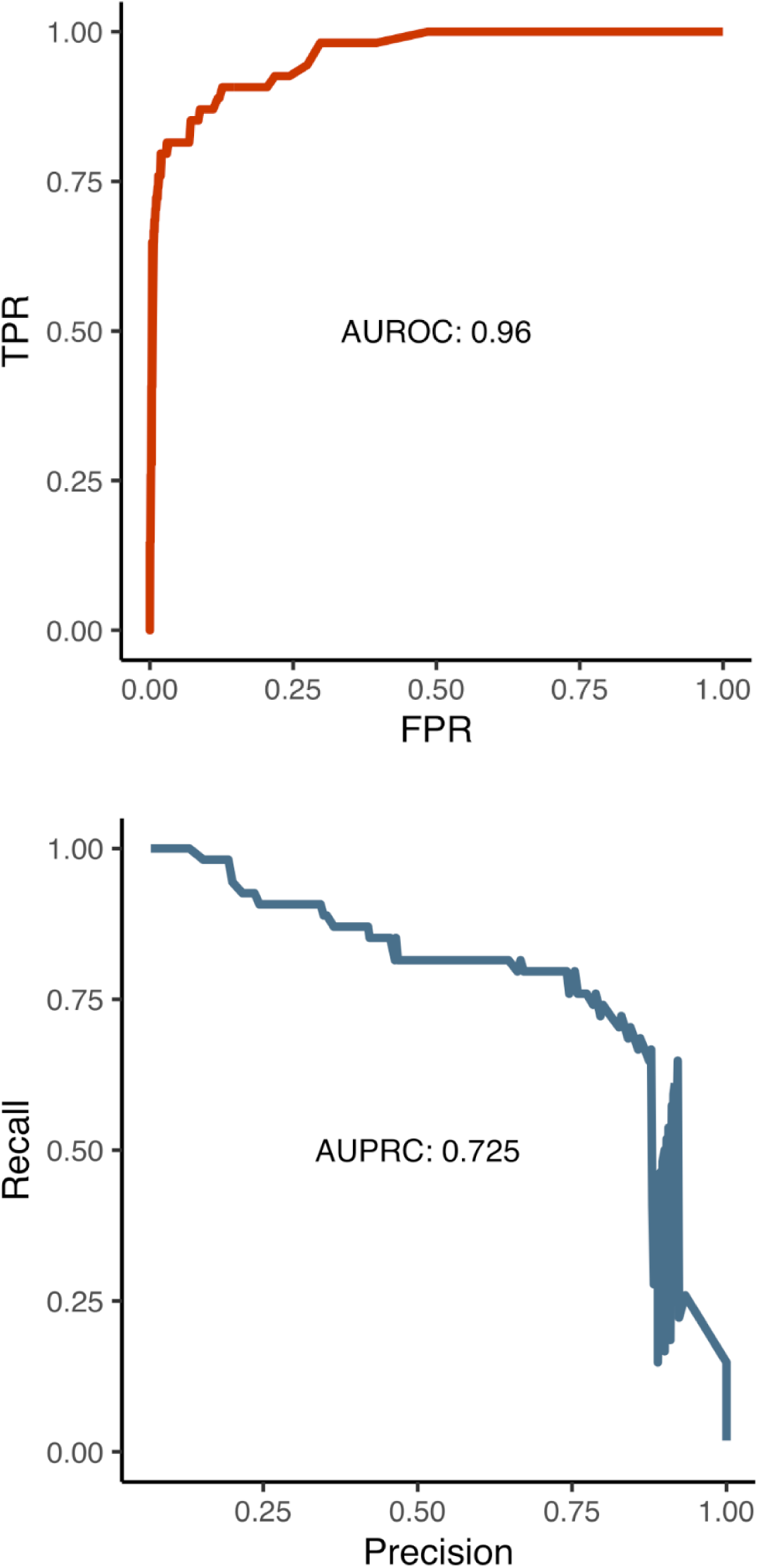
(a) Area under the receiver operating characteristic (AUROC) of our random forest (RF) classifier model. (b) Area under the precision recall curve (AUPRC) of our RF classifier model.

**Figure 3.**
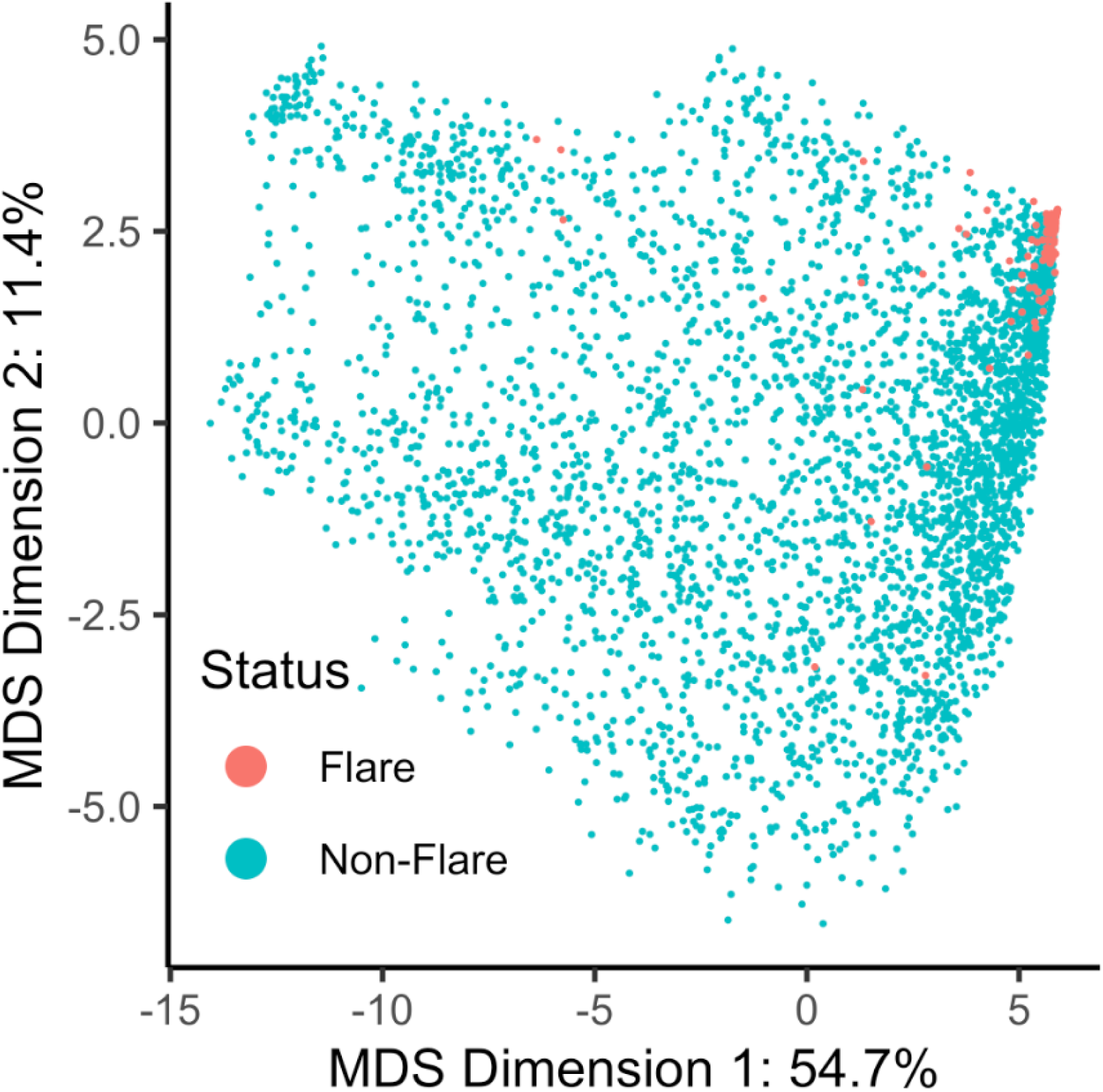
Multidimensional scaling (MDS) plot of all individual ASUC and non-ASUC admissions to visually represent similarity among ASUC admissions. Blue points represent non-ASUC admissions and red numbers represent ASUC admissions. The low pairwise distances between ASUC admissions suggest that claims data are sufficient to detect a high level of similarity amongst ASUC admissions and separate them from non-ASUC admissions.

**Table 2:** Confusion matrix showing the performance of our random forest (RF) model on an internal validation dataset.

|  | **ASUC admission (n=54)** | **Non-ASUC admission (n=743)** |
| --- | --- | --- |
| ASUC admission (Model) | 44 | 26 |
| Non-ASUC admission (Model) | 10 | 717 |

Logistic regression and support vector machine (SVM) models were also trained and tested on our dataset; the RF classifier was found to have the best performance. The logistic regression model had a test AUROC of 0.935; the SVM model had a test AUROC of 0.783. Parameters for each of these models are listed in **Supplementary Table S1**.

Mean decrease in Gini error was computed for the RF model with removal of each feature to estimate feature importance. Based on this relative ranking, the top six most important features in the model were ICD-10 0DBN8ZX (endoscopic diagnostic excision of sigmoid colon), length of stay, age, ICD-10 0DBP8ZX (endoscopic diagnostic excision of rectum), CPT 74020 (radiologic examination of abdomen, complete, including decubitus and/or erect views), and CPT 99151 (moderate sedation services). (**Figure 4**).

**Figure 4:**
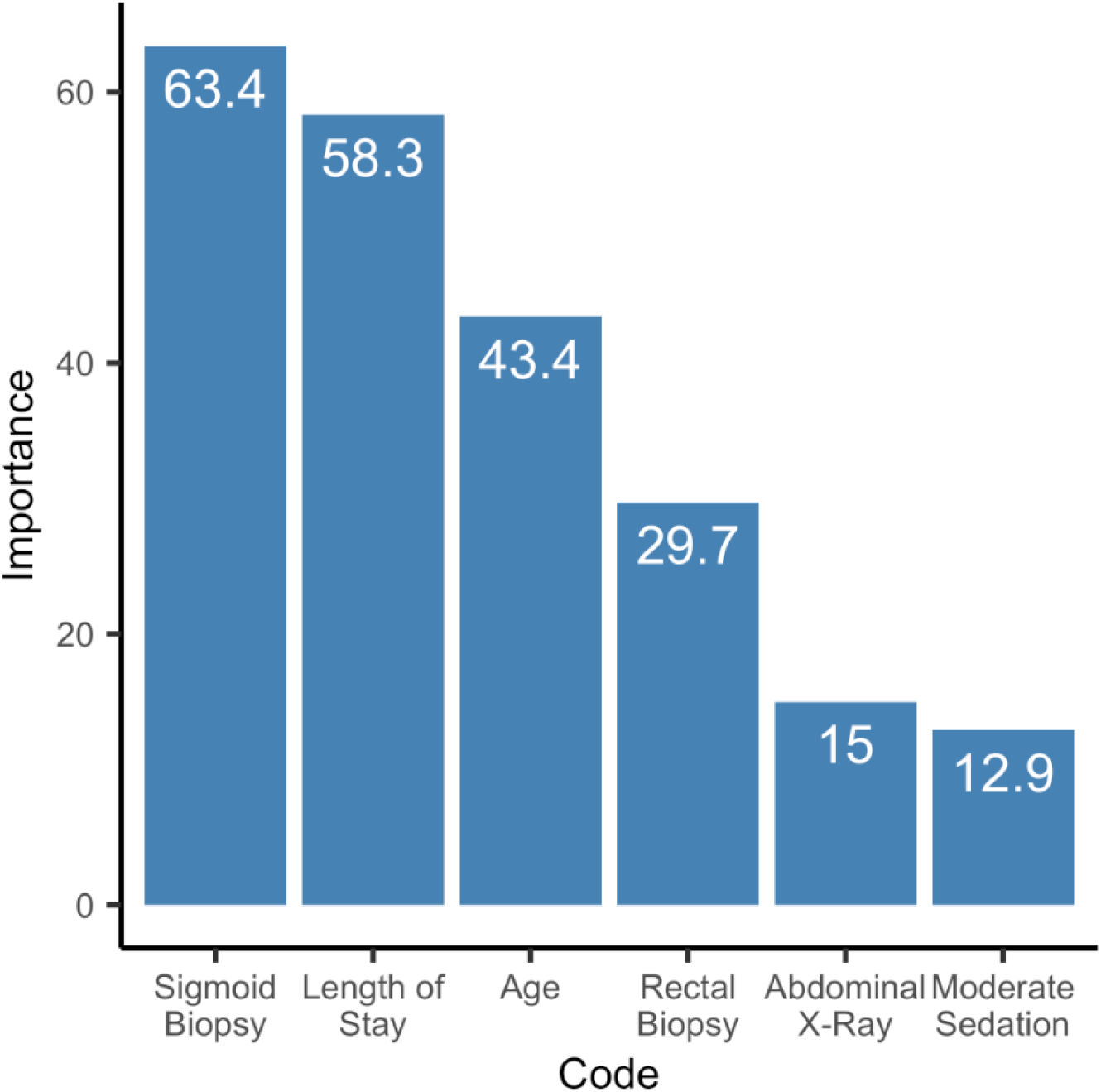
Relative feature importance of each feature included in our random forest (RF) model, measured in mean decrease in Gini error upon removal of this feature from the model.

## Discussion

The granularity of clinical terminologies is an important consideration for many clinical and research activities. In this study, we address this issue by developing a proof-of-concept machine learning-based methodology for automatically identifying patients with un-codeable conditions (i.e., conditions for which there is no unique diagnosis or billing code) that are hidden within structured observational data. In particular, we are able to identify patients with UC who have been admitted for an episode of ASUC versus those who have been admitted for non-ASUC reasons from administrative claims data (diagnosis codes, procedure codes, and drug codes) alone. Our model’s AUROC is 0.96, AUPRC is 0.73, specificity is 96.5%, and negative predictive value (NPV) is 98.6%, suggesting that it is a particularly reliable way to filter out non-ASUC admissions among UC patients from administrative claims data. Its ability to flag true ASUC episodes is also robust: if a hospitalization was flagged by our model, the odds ratio (OR) of it being a true ASUC diagnosis was approximately 46. Upon examining the false positives flagged by our model, 50% of them were of patients who had been admitted for severe UC exacerbations that closely resembled but did not precisely fit ASUC diagnostic criteria. This supports the intended use of our model for identifying clinically significant severe UC exacerbation episodes and generating accurate datasets that can be used to construct cohorts for future retrospective observational research, as these false positive severe exacerbations should likely be included in the final curated dataset anyway. Significant predictors of ASUC admissions from administrative claims codes included endoscopic biopsy of sigmoid colon, length of stay, age, endoscopic biopsy of rectum, abdominal x-ray, and moderate sedation services.

### Comparison to Prior Work

Prior studies that attempt to better understand this condition have struggled to come up with consistent, reliable definitions to identify a UC exacerbation from administrative data. Exacerbations have been defined using codes for colectomy, hospitalizations with an IBD code as a diagnosis, and new steroid prescription codes.^12–14^ Importantly, none of these definitions include endoscopy codes, despite the fact that endoscopic procedures are an essential component of UC exacerbation evaluation and management. Furthermore, the accuracy of these various definitions is unknown; the distribution of true exacerbations versus false positives in their cohorts is unclear. One recent Danish study attempted to identify mild, moderate, and severe exacerbations from administrative data - a slightly different classification task from ours - achieving an AUC of about 0.62-0.67.^19^ This study used fecal calprotectin levels to diagnose true flares, an uncommonly used definition without clear cutoff values.^20^ Our classifier distinguishes ASUC-related hospital admissions from non-ASUC admissions in patients with a history of UC, achieving an AUC of 0.96.

Another prior rules-based classifier uses the codes for steroid administration among UC patients as a surrogate marker for a flare, however the authors acknowledge that there are many other confounding diagnoses that are treated with systemic steroids (e.g., a COPD exacerbation) that make this an imprecise marker for flares. Notably, our model does not find that steroid administration is an important predictor, possibly for the same reason.

### Lack of Granular Clinical Data is Not a Barrier

These findings from our model reassuringly exhibit some consistency with what truly happens in practice at our institution. Patients with a history of UC who are admitted to our institution for ASUC typically undergo a standard admission workup including a flexible endoscopy with biopsies to assess severity of disease and biopsies for cytomegalovirus (CMV), abdominal x-ray imaging to rule out toxic megacolon, infectious stool studies, a serum C-reactive protein (CRP) lab study, and a short course of steroids. Many of these workup components were found to be among the most significant codes in our model. The endoscopic biopsy codes in our model likely represent biopsies to rule out CMV and other conditions. The moderate sedation code in our model likely represents the fact that ASUC patients are typically in significant pain and are unable to tolerate an endoscopic procedure without moderate sedation. Interestingly, our model did not find that steroids were a significant predictor of ASUC admissions. This may be because steroids are nonspecific drugs that are used to treat a number of different conditions unrelated to ASUC as well. Furthermore, steroids are often transiently used empirically in patients with a history of UC, until it is clarified that the patient is not having a true episode of ASUC.

### Limitations

It is important to acknowledge this study’s limitations. Our labeled training dataset of ASUC admissions was generated through chart review, performed by a panel of gastroenterologists at our institution. However, there were no standardized clinical criteria by which the historical gastroenterologist who actually evaluated the patient determined whether the patient was experiencing an ASUC episode or not, especially since the definitions and categorizations of acute UC exacerbations have been changing over time.^5^ This may have introduced some subjectivity into the labeling methodology for our training dataset. Data available in the chart was not granular enough for our panel of gastroenterologists to classify admissions as ASUC or non-ASUC by objective clinical criteria or scoring systems such as the Mayo Score/Disease Activity Index or Truelove and Witts Severity Index.^16,21^ The severity of ASUC episodes were also not graded by our panel of gastroenterologists due to lack of granularity in the chart. However, inpatient admission is likely a reliable surrogate marker of ASUC severity.^5^ Another important limitation is that this is a single-institution study with no external validation of our model, therefore its performance cannot be generalized to other institutions.

Finally, while incorporating additional data sources would certainly improve the predictive performance of the model, our goal was to demonstrate that a dataset of limited elements could accomplish this task reliably to simulate how it would be used in the future on administrative datasets. For example, while labs may have been useful predictors, LOINC codes were deliberately not included in our training data because these codes are typically not part of claims databases.

## Conclusion

In conclusion, there is currently no modality by which certain conditions without their own unique diagnosis or billing code such as ASUC can be rapidly identified from structured observational data in an automated fashion for research or clinical purposes, which may further exacerbate our poor understanding of them. Building tools to identify these conditions can help us better understand them by maximizing our potential to learn from real-world data. In this study, we show that physician-defined characteristics - rather than administrative labels - can be used to identify ASUC from administrative claims data using a machine learning classifier. Specifically, we have developed a machine learning methodology for identifying inpatient admissions for ASUC from administrative claims data alone. This model may be used to label administrative claims databases and automatically generate reasonably accurate datasets of inpatient admissions for ASUC. These datasets may enable further clinically impactful research into the treatment of ASUC, risk factors for ASUC, and the ability to identify patients at risk for an episode of ASUC. Our approach can be applied to rapid large-scale identification of these other conditions as well to enable further study and a better understanding of them.

## Data Availability Statement

The data underlying this article cannot be shared publicly for the privacy of the patients who were included in this study, due to the data containing protected health information on these patients.

## Author Contributions

JSM: study conceptualization, design, analysis, drafting, reviewing, and finalizing manuscript

DC: study conceptualization, design, analysis, drafting, reviewing, and finalizing manuscript

DW: study conceptualization, design, data collection, reviewing and finalizing manuscript

WY: study conceptualization, data analysis, reviewing and finalizing manuscript

MF: study design, data collection, reviewing and finalizing manuscript

CJK: study conceptualization, data analysis, reviewing and finalizing manuscript

YZ: study conceptualization, reviewing and finalizing manuscript

VP: study conceptualization, design, reviewing, and finalizing manuscript

JF: study conceptualization, design, reviewing, and finalizing manuscript

GAB: study conceptualization, design, reviewing, and finalizing manuscript

---

**Supplementary Figure 1:**
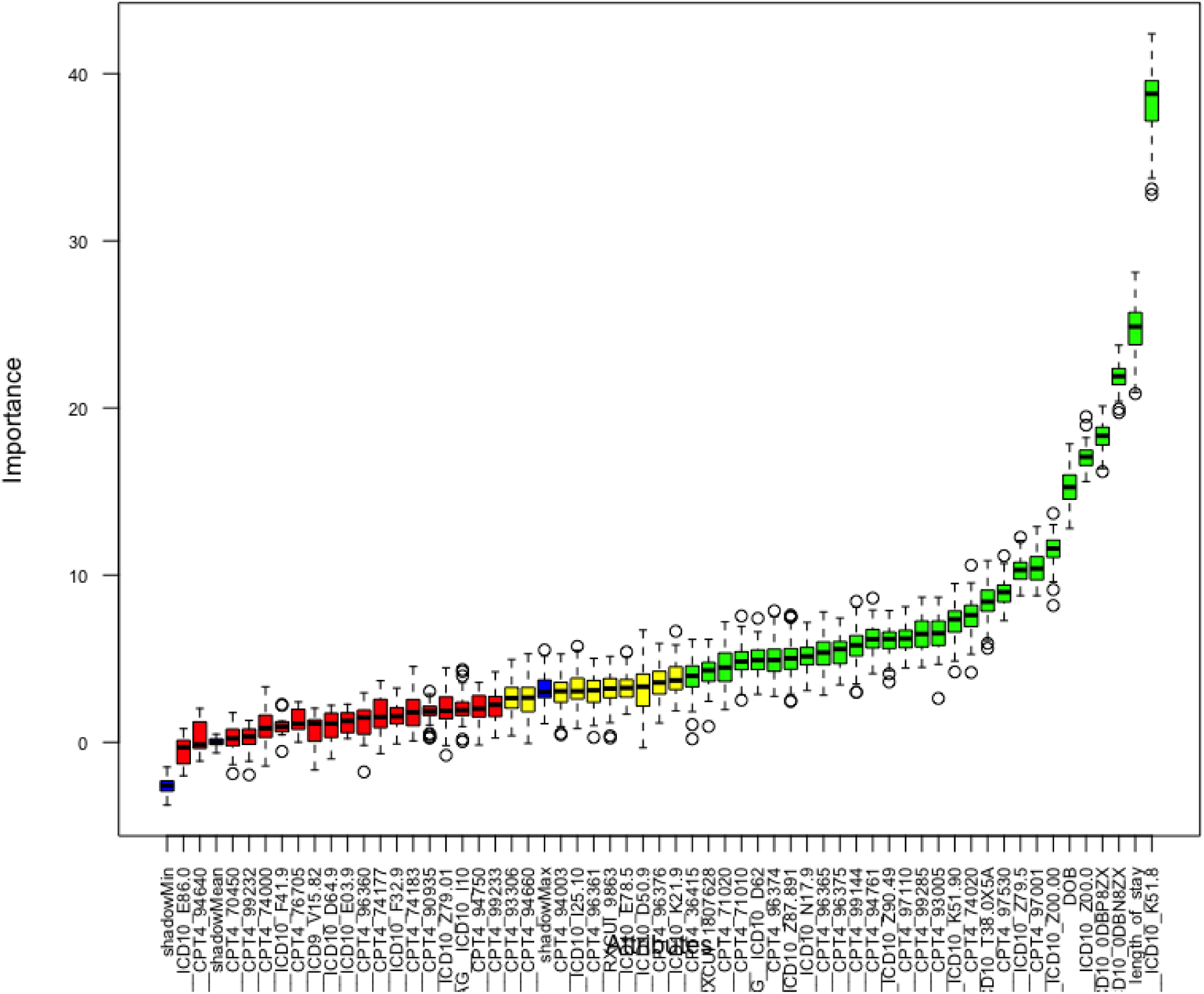
Boruta significant features output. Green are significant, yellow are of moderate significance, red are not significant. Significant (green) features only were included in the final RF model.

**Supplementary Figure 2:**
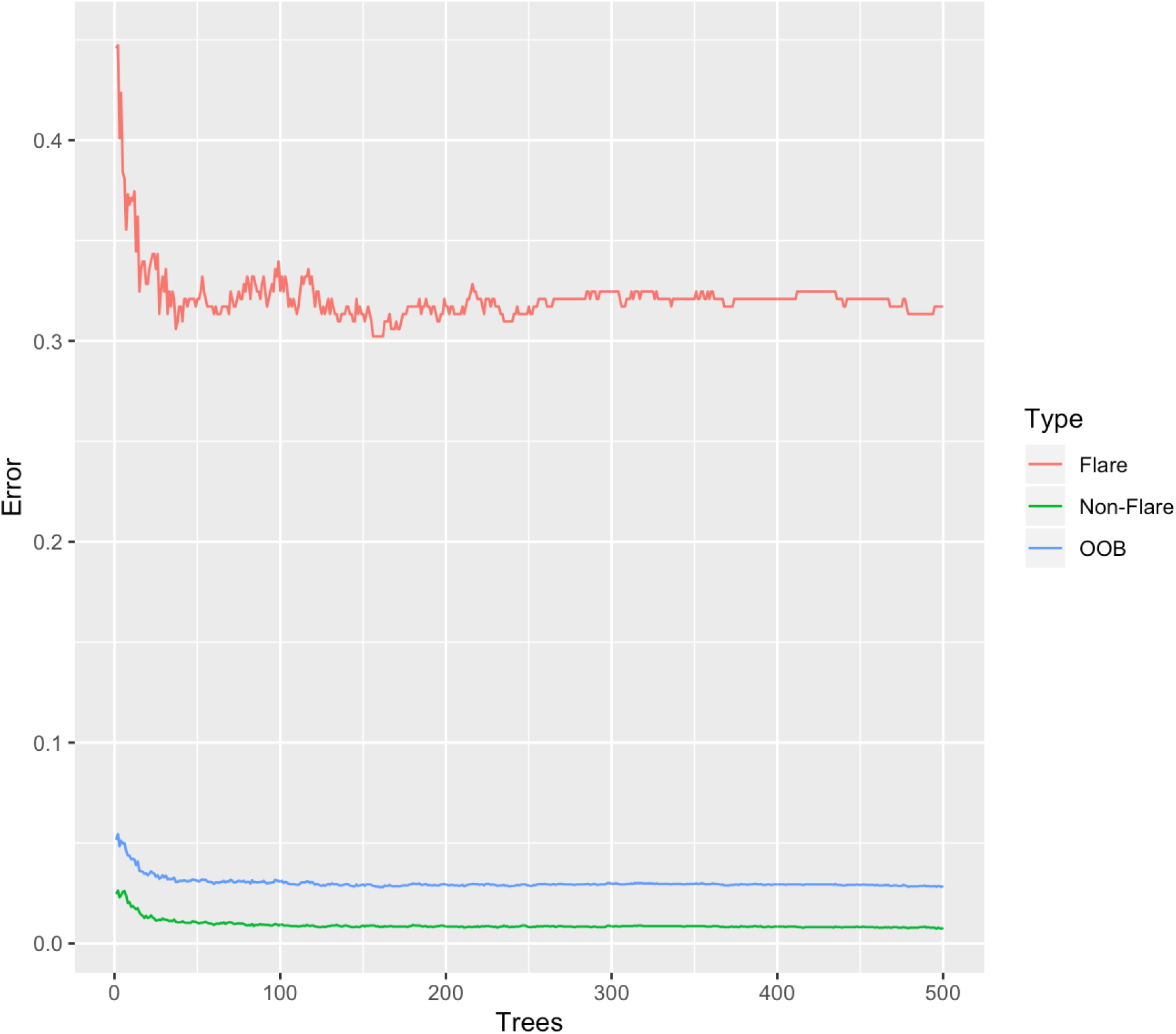
Parameter tuning of the random forest (RF) classifier model. The RF classifier out-of-bag (OOB) error stabilized at approximately 100 trees.

**Supplementary Table 1:** TRIPOD reporting criteria for our study.

| Section/Topic | Item | Checklist Item | Page |
| --- | --- | --- | --- |
| **Title and abstract** |  |  |  |
| Title | 1 | Identify the study as developing and/or validating a multivariable prediction model, the target population, and the outcome to be predicted. | 1 |
| Abstract | 2 | Provide a summary of objectives, study design, setting, participants, sample size, predictors, outcome, statistical analysis, results, and conclusions. | 3-4 |
| **Introduction** |  |  |  |
| Background and objectives | 3a | Explain the medical context (including whether diagnostic or prognostic) and rationale for developing or validating the multivariable prediction model, including references to existing models. | 7 |
|  | 3b | Specify the objectives, including whether the study describes the development or validation of the model or both. | 7 |
| **Methods** |  |  |  |
| Source of data | 4a | Describe the study design or source of data (e.g., randomized trial, cohort, or registry data), separately for the development and validation data sets, if applicable. | 7-8 |
|  | 4b | Specify the key study dates, including start of accrual; end of accrual; and, if applicable, end of follow-up. | 7 |
| Participants | 5a | Specify key elements of the study setting (e.g., primary care, secondary care, general population) including number and location of centres. | 7-8 |
|  | 5b | Describe eligibility criteria for participants. | 8 |
|  | 5c | Give details of treatments received, if relevant. | N/A |
| Outcome | 6a | Clearly define the outcome that is predicted by the prediction model, including how and when assessed. | 9-10 |
|  | 6b | Report any actions to blind assessment of the outcome to be predicted. | N/A |
| Predictors | 7a | Clearly define all predictors used in developing or validating the multivariable prediction model, including how and when they were measured. | 8-9 |
|  | 7b | Report any actions to blind assessment of predictors for the outcome and other predictors. | N/A |
| Sample size | 8 | Explain how the study size was arrived at. | 7-8 |
| Missing data | 9 | Describe how missing data were handled (e.g., complete-case analysis, single imputation, multiple imputation) with details of any imputation method. | N/A |
| Statistical analysis methods | 10a | Describe how predictors were handled in the analyses. | 10-11 |
|  | 10b | Specify type of model, all model-building procedures (including any predictor selection), and method for internal validation. | 10-11 |
|  | 10d | Specify all measures used to assess model performance and, if relevant, to compare multiple models. | 10-11 |
| Risk groups | 11 | Provide details on how risk groups were created, if done. | 9-10 |
| **Results** |  |  |  |
| Participants | 13a | Describe the flow of participants through the study, including the number of participants with and without the outcome and, if applicable, a summary of the follow-up time. A diagram may be helpful. | 24 |
|  | 13b | Describe the characteristics of the participants (basic demographics, clinical features, available predictors), including the number of participants with missing data for predictors and outcome. | 23 |
| Model development | 14a | Specify the number of participants and outcome events in each analysis. | 23 |
|  | 14b | If done, report the unadjusted association between each candidate predictor and outcome. | Not done |
| Model specification | 15a | Present the full prediction model to allow predictions for individuals (i.e., all regression coefficients, and model intercept or baseline survival at a given time point). | 28 |
|  | 15b | Explain how to use the prediction model. | 19 |
| Model performance | 16 | Report performance measures (with CIs) for the prediction model. | 13 |
| **Discussion** |  |  |  |
| Limitations | 18 | Discuss any limitations of the study (such as nonrepresentative sample, few events per predictor, missing data). | 17-18 |
| Interpretation | 19b | Give an overall interpretation of the results, considering objectives, limitations, and results from similar studies, and other relevant evidence. | 19 |
| Implications | 20 | Discuss the potential clinical use of the model and implications for future research. | 19 |
| **Other information** |  |  |  |
| Supplementary information | 21 | Provide information about the availability of supplementary resources, such as study protocol, Web calculator, and data sets. | 29-32 |
| Funding | 22 | Give the source of funding and the role of the funders for the present study. | 19 |

We recommend using the TRIPOD Checklist in conjunction with the TRIPOD Explanation and Elaboration document.

**Supplementary Table 2:** Parameters for the logistic regression (LR), random forest (RF), and support vector machine (SVM) models trained on the training dataset.

| **Model** | **Parameter** | **Value** |
| --- | --- | --- |
| Logistic regression | No. Fisher scoring iterations | 20 |
| Random forest | No. variables randomly sampled at each split | 4 |
|  | No. trees | 300 |
| Support vector machine | Type | C-classification |
|  | Kernel | Linear |
|  | Cost of constraints violation | 1 |
|  | No. support vectors | 346 |

## References

1. ICD - ICD-10-CM - International Classification of Diseases, Tenth Revision, Clinical Modification. June 28, 2021. Accessed July 1, 2021. https://www.cdc.gov/nchs/icd/icd10cm.htm

2. ICD Coding for Rare Diseases. Accessed July 1, 2021. https://rarediseases.info.nih.gov/guides/pages/123/icd-coding-for-rare-diseases

3. Aymé S, Bellet B, Rath A. Rare diseases in ICD11: making rare diseases visible in health information systems through appropriate coding. Orphanet J Rare Dis. 2015;10(1):1–14.

4. Zghebi SS, Mamas MA, Ashcroft DM, et al. Development and validation of the DIabetes Severity SCOre (DISSCO) in 139 626 individuals with type 2 diabetes: a retrospective cohort study. BMJ Open Diab Res Care. 2020;8(1). doi:10.1136/bmjdrc-2019-000962

5. Feuerstein JD, Isaacs KL, Schneider Y, et al. AGA Clinical Practice Guidelines on the Management of Moderate to Severe Ulcerative Colitis. Gastroenterology. 2020;158(5):1450–1461.

6. Gallo G, Kotze PG, Spinelli A. Surgery in ulcerative colitis: When? How? Best Pract Res Clin Gastroenterol. 2018;32–33. doi:10.1016/j.bpg.2018.05.017

7. Narula N, Marshall JK, Colombel JF, et al. Systematic Review and Meta-Analysis: Infliximab or Cyclosporine as Rescue Therapy in Patients With Severe Ulcerative Colitis Refractory to Steroids. Am J Gastroenterol. 2016;111(4). doi:10.1038/ajg.2016.7

8. Collins P, Rhodes J. Ulcerative colitis: diagnosis and management. BMJ. 2006;333(7563):340–343.

9. Walmsley RS, Ayres RCS, Pounder RE, Allan RN. A simple clinical colitis activity index. Gut. 1998;43(1):29–32. doi:10.1136/gut.43.1.29

10. Powell-Tuck J, Bown RL, Lennard-Jones JE. A comparison of oral prednisolone given as single or multiple daily doses for active proctocolitis. Scand J Gastroenterol. 1978;13(7):833–837.

11. Lewis JD, Chuai S, Nessel L, Lichtenstein GR, Aberra FN, Ellenberg JH. Use of the noninvasive components of the Mayo score to assess clinical response in ulcerative colitis. Inflamm Bowel Dis. 2008;14(12):1660–1666.

12. Lund JL, Stürmer T, Porter CQ, Sandler RS, Kappelman MD. Thiazolidinedione use and ulcerative colitis-related flares: an exploratory analysis of administrative data. Inflamm Bowel Dis. 2011;17(3):787–794.

13. Kristensen SL, Ahlehoff O, Lindhardsen J, et al. Disease activity in inflammatory bowel disease is associated with increased risk of myocardial infarction, stroke and cardiovascular death--a Danish nationwide cohort study. PLoS One. 2013;8(2):e56944.

14. Lewis JD, Aberra FN, Lichtenstein GR, Bilker WB, Brensinger C, Strom BL. Seasonal variation in flares of inflammatory bowel disease. Gastroenterology. 2004;126(3):665–673.

15. Lewis JD, Brensinger C, Bilker WB, Strom BL. Validity and completeness of the General Practice Research Database for studies of inflammatory bowel disease. Pharmacoepidemiology and drug safety. 2002;11(3). doi:10.1002/pds.698

16. Truelove SC, Witts LJ. Cortisone in ulcerative colitis; final report on a therapeutic trial. Br Med J. 1955;2(4947):1041–1048.

17. Kursa MB, Rudnicki WR. Feature Selection with theBorutaPackage. J Stat Softw. 2010;36(11). doi:10.18637/jss.v036.i11

18. Collins GS, Reitsma JB, Altman DG, Moons KG. Transparent Reporting of a multivariable prediction model for Individual Prognosis or Diagnosis (TRIPOD): the TRIPOD statement. Ann Intern Med. 2015;162(1). doi:10.7326/M14-0697

19. Burisch J, Zhang H, Choong CK, et al. Validation of claims-based indicators used to identify flare-ups in inflammatory bowel disease. Therap Adv Gastroenterol. 2021;14. doi:10.1177/17562848211004841

20. Rubin DT, Ananthakrishnan AN, Siegel CA, Sauer BG, Long. ACG Clinical Guideline: Ulcerative Colitis in Adults. Am J Gastroenterol. 2019;114(3). doi:10.14309/ajg.0000000000000152

21. Schroeder KW, Tremaine WJ, Ilstrup DM. Coated oral 5-aminosalicylic acid therapy for mildly to moderately active ulcerative colitis. A randomized study. N Engl J Med. 1987;317(26):1625–1629.

